# A Thermostable Cas12b from *Brevibacillus* Leverages One-pot Detection of SARS-CoV-2 Variants of Concern

**DOI:** 10.1101/2021.10.15.21265066

**Authors:** Long T. Nguyen, Nicolas C. Macaluso, Brianna L.M. Pizzano, Melanie N. Cash, Jan Spacek, Jan Karasek, Rhoel R. Dinglasan, Marco Salemi, Piyush K. Jain

## Abstract

Current SARS-CoV-2 detection platforms lack the ability to differentiate among variants of concern (VOCs) in an efficient manner. CRISPR (Clustered Regularly Interspaced Short Palindromic Repeats) has the potential to transform diagnostics due to its programmability. However, many of the CRISPR-based detection methods are reliant on either a multi-step process involving amplification or elaborate guide RNA designs. A complete one-pot detection reaction using alternative Cas effector endonucleases has been proposed to overcome these challenges. Yet, current approaches using *Alicyclobacillus acidiphilus* Cas12b (AapCas12b) are limited by its thermal instability at optimum reverse transcription loop-mediated isothermal amplification (RT-LAMP) reaction temperatures. Herein, we demonstrate that a novel Cas12b from *Brevibacillus* sp. SYP-B805 (referred to as BrCas12b) has robust trans-cleavage activity at ideal RT-LAMP conditions. A competitive profiling study of BrCas12b against Cas12b homologs from other bacteria genera underscores the potential of BrCas12b in the development of new diagnostics. As a proof-of-concept, we incorporated BrCas12b into an RT-LAMP-mediated one-pot reaction system, coined CRISPR-SPADE (CRISPR Single Pot Assay for Detecting Emerging VOCs) to enable rapid, differential detection of SARS-CoV-2 VOCs, including Alpha (B.1.1.7), Beta (B.1.351), Gamma (P.1), and Delta (B.1.617.2) in 205 clinical samples. Notably, a BrCas12b detection signal was observed within 1-3 minutes of amplification, achieving an overall 98.1% specificity, 91.2% accuracy, and 88.1% sensitivity within 30 minutes. Significantly, for samples with high viral load (C_t_ value ≤ 30), 100% accuracy and sensitivity were attained. To facilitate dissemination and global implementation of the assay, we combined the lyophilized one-pot reagents with a portable multiplexing device capable of interpreting fluorescence signals at a fraction of the cost of a qPCR system. With relaxed design requirements, one-pot detection, and simple instrumentation, this assay has the capability to advance future diagnostics.

## MAIN

Since the beginning of the COVID-19 pandemic, many strategies have been explored to develop rapid and sensitive detection kits to drive the diagnostics towards home-based testing^1-5^. Current gold-standard Reverse Transcription - quantitative Polymerase Chain Reaction (RT-qPCR) tests require sophisticated instrumentation as well as intensive labor training, and therefore become a major hurdle for deployment in remote areas. CRISPR-based detection technologies hold promise for future rapid point-of-care diagnosis of infectious diseases and cancer^6,7^. This is in part due to their versatility in implementation and design that lie in the programmability of the guide RNA sequence. Taking advantage of the collateral cleavage property, many Cas effectors have been repurposed for nucleic acid detection such as Cas12a in DETECTR (DNA Endonuclease-Targeted CRISPR Trans Reporter)^5,8^ and in HOLMESv1 (a one-HOur Low-cost Multipurpose highly Efficient System)^9^, Cas12b in HOLMESv2^10^, and Cas13a in SHERLOCK (Specific High Sensitivity Enzymatic Reporter UnLOCKing)^11,12^. Many of these tests employ a two-pot assay in which a pre-amplification step such as RPA (reverse transcription recombinase polymerase amplification) or RT-LAMP (reverse transcription loop-mediated isothermal amplification) is required prior to CRISPR detection. This strategy increases reaction time and chances of carryover contamination. Efforts into developing rapid amplification-free CRISPR tests have been achieved, such as FIND-IT (Fast Integrated Nuclease Detection In Tandem) that supplements Csm6 protein into a Cas13a reaction to trigger a cascade of amplified fluorescence signal^13^. However, these tests rely on guide RNA targeting multiple regions of the SARS-CoV-2 RNA genome, which can be difficult to design to discriminate the variants of concerns (VOCs) due to the low number of mutations. Having the ability to differentiate the VOCs is still dependent on the enrichment of target viral genomes. STOPCovid technology can potentially overcome these challenges by employing a thermostable Cas12b derived from *Alicyclobacillus acidiphilus* (AapCas12b) that can be combined with RT-LAMP into a one-pot detection assay^1^. One major disadvantage of STOPCovid is that the wild-type AapCas12b collateral activity ceases to work above 60°C and requires additional additives, such as taurine, and a longer incubation time to perform robustly due to suboptimal temperature conditions, leading to slower diagnostics. Additionally, the protein’s performance at this temperature range restricts LAMP primer designs since RT-LAMP reactions are typically optimized at 60°C – 65°C^14,15^.

Here, we report the development of a complete one-pot RT-LAMP-coupled CRISPR detection reaction using a novel Cas12b derived from unclassified *Brevibacillus* sp. SYP-B805 (GenBank ID : WP_165214399.1) named BrCas12b. BrCas12b exhibits phenomenal stability at high temperatures (up to 70°C in optimal buffers) which is suitable for coupling with an RT-LAMP reaction. Notably, BrCas12b is shown to have high collateral cleavage (referred to as trans-cleavage) up to the temperature of 64°C without the need for supplemental additives. In addition, BrCas12b works robustly in isothermal amplification buffer, which is an ideal scenario for its incorporation into a complete one-pot reaction. The one-pot RT-LAMP-coupled BrCas12b reaction provides two detection checkpoints: 1) amplification by RT-LAMP that can be tracked by SYTO™ dye and 2) BrCas12b:sgRNA complex detecting amplified targets that can emit a different signal by a Fluorescence Resonance Energy Transfer (FRET)-based reporter. This dual-checkpoint one-pot assay provides a highly accurate level of nucleic acid detection. The broad versatility along with the high specificity of BrCas12b enables us to detect SARS-CoV-2 and distinguish its variants of concerns Alpha (B.1.1.7), Beta (B.1.352), Gamma (P1), and Delta (B.1.617.2). Finally, we develop two low-cost detection methods for fluorescence visualization. The first one requires a color-filtered lens attached to a mobile phone camera and a handheld flashlight. By engaging the flashlight, samples can be detected in the dark by the naked eye or enhanced and magnified through the use of filters and the camera. The second method utilizes a homemade, portable and inexpensive prototype that allows for dual checkpoints of amplification and BrCas12b trans-cleavage activity using FITC and Cy5 channels.

Tian et al. first reported and characterized the BrCas12b from a hot spring bacterium *Brevibacillus* sp. SYP-B805 that exerts high enzymatic target cleavage (cis-cleavage) activity at elevated temperature (up to 65.5°C); however, the trans-cleavage activity remained to be investigated^16^. BrCas12b forms a complex with crRNA and tracrRNA (∼ 130 nt) to recognize and cleave dsDNA target containing a TTN PAM sequence upstream of the complementary binding site (Fig. 1a). We sought to verify the reported BrCas12b cis-cleavage activity by carrying out a temperature-dependent in vitro cleavage assay compared to Cas12b from *Alicyclobacillus acidoterrestris* (AacCas12b) and *Alicyclobacillus acidiphilus* (AapCas12b), two thermostable effector endonucleases with considerable cleavage activity as reported in STOPCovid (Supplementary Fig. 1). A more restrictive PAM sequence TTTN was selected to investigate these three effectors’ performance. Corroborating the study by Tian et al., BrCas12b showed cleavage up to 70°C in Bovine Serum Albumin-containing buffer compared to 52°C and 59°C for AacCas12b and AapCas12b, respectively (Fig. 1b). Thermal stability for AacCas12b, AapCas12b, and BrCas12b was confirmed by differential scanning fluorimetry where the melting temperatures were found to be 55.3°C, 58.3°C, and 63.4°C, respectively (Fig. 1c and Supplementary Fig. 2). Fascinated by the robust performance of BrCas12b, we proceeded to test the effector’s trans-cleavage activity to determine if it is suitable for a one-pot RT-LAMP-coupled detection reaction. As shown by trans-cleavage kinetic analysis, we observed a turnover number of 14.2 s^-1^ and catalytic efficiency of 1.74×10^7^ (M^-1^s^-1^) at 62°C, indicating fast enzymatic activity (Fig. 1d and Supplementary Fig. 3).

**Figure 1.**
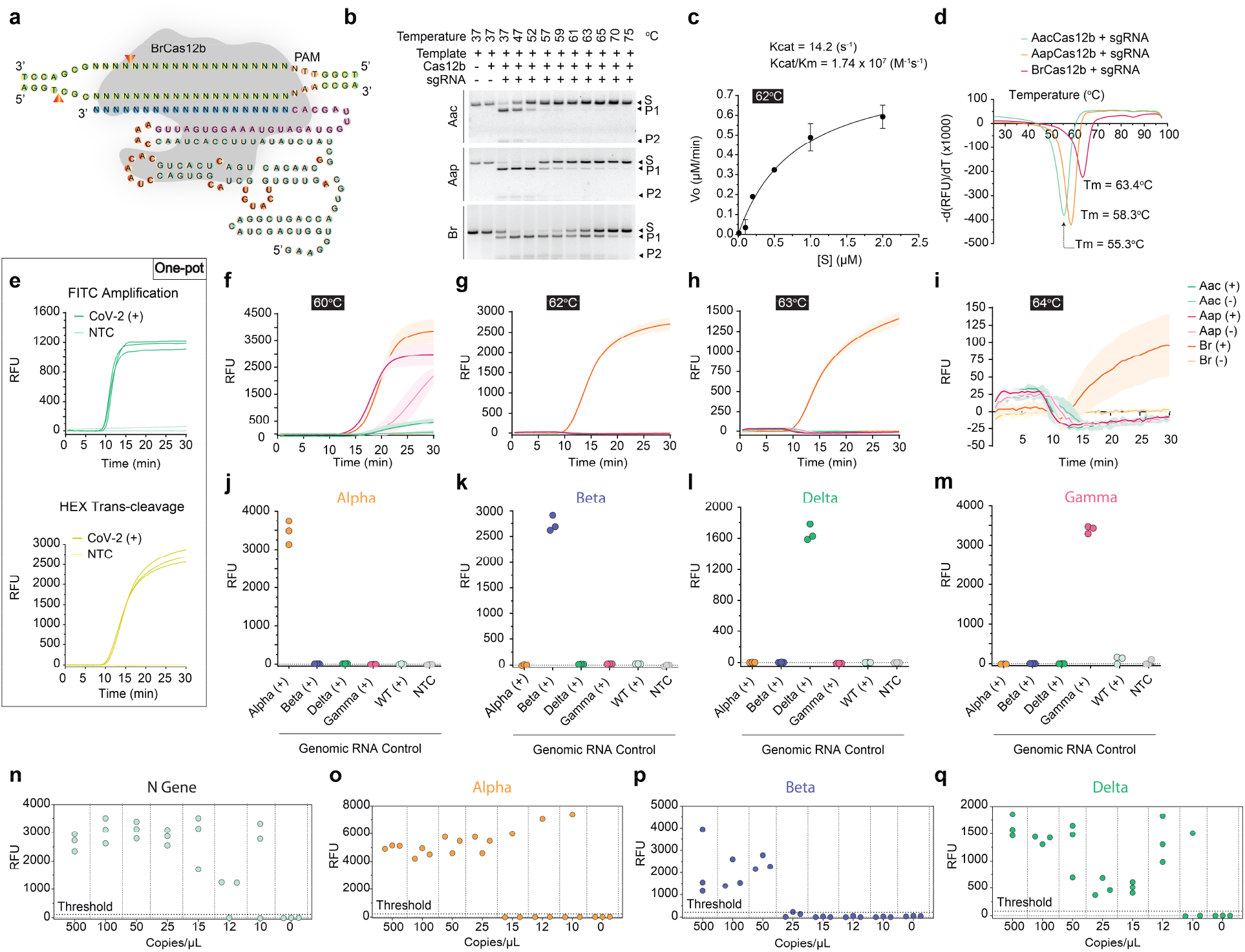
BrCas12b characterization and one-pot specificity & sensitivity testing. (**a**) Schematic of binary complex illustrating cleavage pattern of dsDNA target. (**b**) Temperature-dependent in-vitro cleavage assay of AacCas12b, AapCas12b, and BrCas12b targeting more restrictive PAM TTTG. 125 nM sgRNA:100 nM Cas12b:7 nM dsDNA was combined in 1X NEBuffer 2.1. The experiment was repeated (n = 2) with similar results. (**c**) Michaelis-Menten Kinetics of BrCas12b trans-cleavage at 62°C. (**d**) Differential scanning fluorimetry of AacCas12b, AapCas12b, and BrCas12b complexes. **(e)** Multiplexing using FITC to detect LAMP amplification and HEX to monitor trans-cleavage of BrCas12b. (**f**) – (**i**) Detection capability of BrCas12b via trans-cleavage against AacCas12b and AapCas12b at various temperatures. Fluorescence kinetics within 30 minutes was monitored using a HEX-based reporter. Shaded regions represent standard deviation (n = 3 replicates). STOPCovid LAMP primers were used in (f) to compare performance with AapCas12b. A different set of LAMP primers (DETECTR) with optimal performance at a temperature higher than 60°C were used in (g), (h), and (i). (+) denotes the presence of the SARS-CoV-2 genomic RNA control, and (-) signifies the non-template control (NTC). (**j**) – (**m**) Specificity testing using a pair of sgRNA and LAMP primers targeting SARS-CoV-2 variants (n = 3 replicates). (**n**) – (**q**) Limit of detection on N gene and S gene targeting alpha (B.1.1.7), beta (B.1.352), and delta (B.1.617.2), respectively (n = 3 replicates).

The above observations provide advantages when coupling with an RT-LAMP reaction for detection purposes. First, the high optimum temperature of BrCas12b lies within the range of ideal RT-LAMP operating conditions. Additionally, we noticed that BrCas12b performed well in the isothermal amplification buffer used in the RT-LAMP reaction. After iterative optimization using many common additives such as proline, taurine, BSA, and betaine, we did not observe an enhancement in the trans-cleavage activity (Supplementary Fig. 4). Therefore, we formulated an RT-LAMP-coupled BrCas12b reaction in the absence of additives. By multiplexing the assay with two different reporters, we were able to track both target amplification and BrCas12b trans-cleavage activity in a one-pot reaction. This allowed us to confirm the presence of nucleic acid targets via the two checkpoints. Since the LAMP reaction is prone to non-specific primer-dimer amplification, the BrCas12b trans-cleavage activity serves as a final verdict for detection (Fig. 1e). We observed that the trans-cleavage activity of BrCas12b has a 1–3-minute delay after amplification, indicating minimal inhibition by the LAMP reaction on activity. Multiple studies have shown that when combining RT-LAMP/RPA with CRISPR reaction, the sensitivity of detection is reduced significantly possibly due to operating temperature differences, an inhibitory effect of an excessive amount of amplified product, differences in optimal buffer and salt conditions, and unwanted nonspecific trans-cleavage of Cas effector on primers^1,13,17-19^. However, having observed such a robust trans-cleavage with a small delay after amplification, we consider that BrCas12b has circumvented many of these issues.

We then systematically tested the one-pot reaction at various temperatures ranging from 60°C to 64°C to evaluate detection performance among Cas12b. To compare with STOPCovid, we tested its LAMP primers at 60°C but with a higher concentration of Cas protein and sgRNA. We selected the LAMP primers used in DETECTR for the remaining temperatures because STOPCovid primers are not optimal above 60°C (Fig. 1f-i)^5^. BrCas12b was observed to work robustly up to 63°C and with reduced activity at 64°C, whereas, as expected, AacCas12b and AapCas12b failed at temperatures above 60°C (Fig. 1f-i). Additionally, false positives were occasionally observed when STOPCovid LAMP primers and high concentrations of AapCas12b and sgRNA were used (Fig. 1f).

The thermal stability of BrCas12b enabled us to design LAMP primers with less restrictive parameters compared to STOPCovid to comprehensively distinguish SARS-CoV-2 VOCs from its original Wuhan strain. We systematically tested each variant including Alpha (B.1.1.7), Beta (B.1.351), Gamma (P1), and Delta (B.1.6127.2) against one another and the Wuhan strain (Supplementary Fig. 5). The pair of LAMP primers and sgRNA targeting each variant exhibited high specificity with no relative cross-target detection (Fig. 1j-m). For sensitivity testing, we sought to determine the limit of detection (LoD) of nucleocapsid gene (N) for the presence of SARS-CoV-2 virions and non-N mutated genes for the prevalent VOCs. The assay confirmed the estimated LoD of 15 copies/µL and 25 copies/µL, 50 copies/µL, and 12 copies/µL for N gene, Alpha, Beta, and Delta, respectively (Fig. 1n,o).

We hereafter refer to the RT-LAMP-coupled BrCas12b test as CRISPR-SPADE (**S**ingle **P**ot **A**ssay for **D**etecting **E**merging VOCs). We proceeded to validate CRISPR-SPADE in clinical samples including 57 Delta positive (B.1.617.2), 33 Alpha positive (B.1.1.7), 17 Gamma positive (P.1), 1 Beta positive (B.1.351), 44 Wuhan SARS-CoV-2, and 53 negative samples. N gene and non-N genes were tested in parallel. The detection of the N gene served as a basis for the presence of SARS-CoV-2, whereas the non-N genes were used to differentiate the variants. An 88.1 % sensitivity in the N gene for all samples was reached; likewise, a 93.9%, 94.6%, and 76.5% sensitivity were achieved after 30 minutes in the non-N gene for the alpha, delta, and gamma samples, respectively (Fig. 2.a-c). LAMP primers play a crucial role in the sensitivity and specificity of the BrCas12b one-pot detection (Fig. 2d). Quality in LAMP primer design can aid in the reduction of primer-dimer amplification^14^. During the development of the one-pot detection reaction, we occasionally observed non-specific amplification, which was the main cause for false negatives in low copy samples due to the consumption of resources. The high optimum temperature of BrCas12b alleviates some of the LAMP primer design restrictions and thus allows for more flexibility in primer selection. Specifically, in samples with a C_t_ value less than 30, 100% sensitivity was accomplished, and in samples above a C_t_ value of 30, we observed a reduced performance in sensitivity (Fig. 2e). With negative samples considered, we achieved an overall 91.2% accuracy and 98.1 % specificity with one false positive detected out of a total of 205 samples (Supplementary Fig. 6,7). Patient samples detected with variants were confirmed through next-generation sequencing (Supplementary Fig. 8).

**Figure 2.**
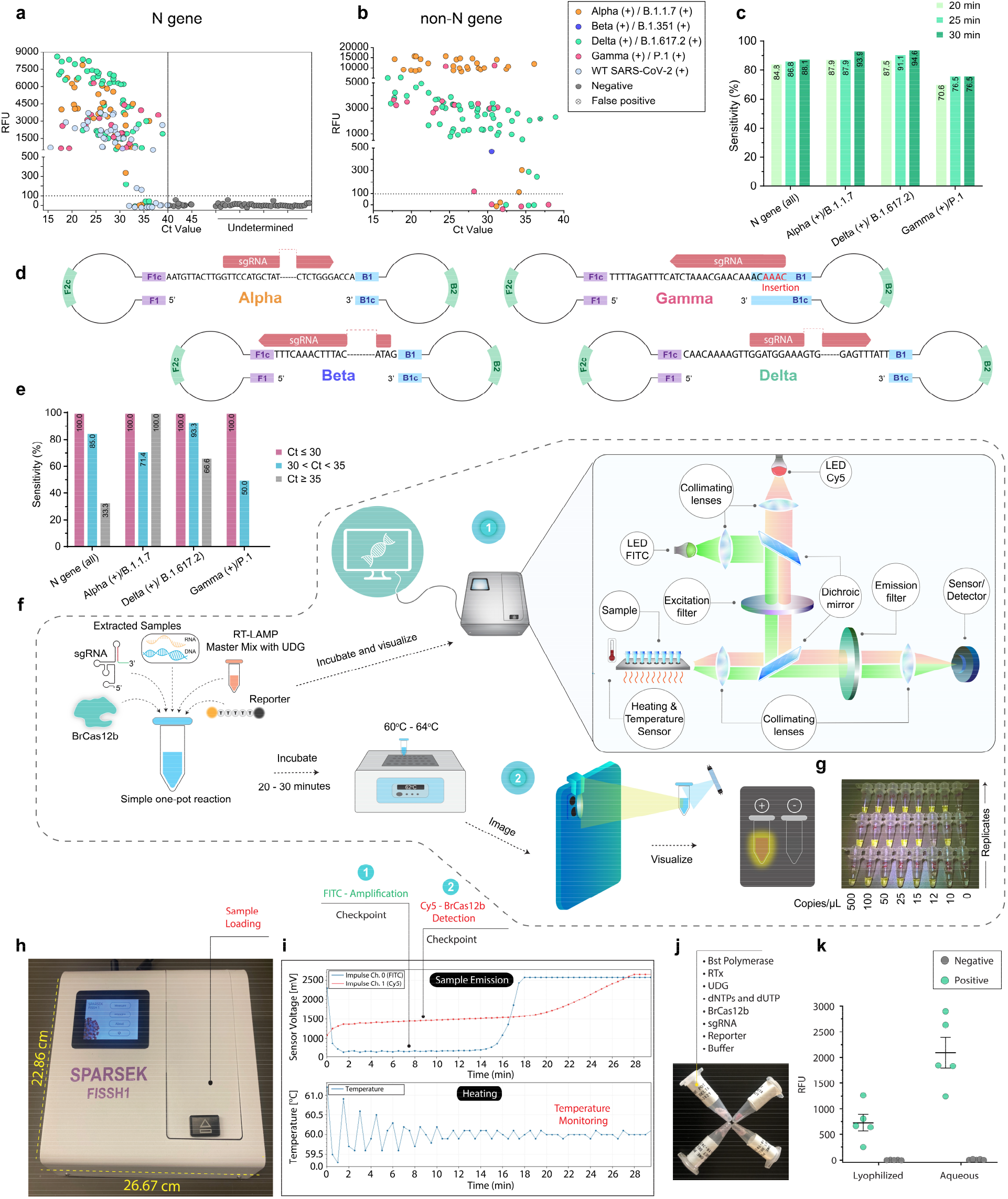
Clinical validation of one-pot BrCa12b detection assay and portable diagnostic systems. (**a**) and (**b**) One-pot patient sample detection. The N gene target indicates the presence of SARS-CoV-2 while non-N gene targets indicate detection of variants. Fluorescence measurements were taken at t = 30 minutes. (**c**) Percent accuracy of the one-pot detection at 20-, 25-, and 30-minutes targeting variants and WT SARS-CoV-2. (**d**) Self-hybridizing loop structure acts as a seed for exponential LAMP amplification which subsequently serves as a basis for BrCas12b one-pot detection of VOCs. Guide RNA designed to target the dumbbell region of the LAMP products are shown. (**e**) Percent accuracy with respect to Ct value for SARS-CoV-2 variant detection.(**f**) One-pot assembly and comparison of a portable in-house detection instrument FISSH) and an at-home detection method using a mobile phone with a simple lens. (**g**) depiction of an image taken by the mobile phone in (e). (**h**) and (**i**) FISSH footprint and dual-channel graph generated from a positive sample. (**j**) representation of lyophilized one-pot detection reaction. (**k**) Lyophilized samples compared to the standard one-pot reaction. Fluorescence measurements were taken at t = 30 minutes and the Mean±SD (n = 5) are indicated.

We further developed two methods of fluorescence-based detection: an inexpensive lens attached to a mobile phone camera, and a multiplexing detection prototype engineered to track RT-LAMP amplification and BrCas12b trans-cleavage simultaneously (Fig. 2f-i). The phone-based detection activates via a UVA-blue flashlight (410 nm – 415 nm) and a combination of low-cost yellow and orange filtered lenses with a clip (cost < $5). This combined lens system allowed for the detection of FRET-based HEX reporters in a dark setting (Fig. 2f). We envision that this approach, with proper safety precautions, could be used for home-based testing, as demonstrated by others^3,4,13,20^. The portable multiplexing detection prototype was built with two wavelengths in the optical system: a LED working at 490/26 nm excitation for FITC, and a second LED working at 625/17 nm with the same length of impulse for Cy5 excitation. A combination of reporter pairs could be used for this portable device such as SYTO™ 62 using Cy5 channel for tracking amplification and FRET-based FAM reporter using the FITC channel for detection of BrCas12b trans-cleavage activity, or the SYTO™ 9 and Cy5 reporter pair for the opposite channels. This prototype came with a built-in touch screen for ease of use (Fig. 2g) and could potentially be programmed to connect to a mobile phone or a tablet via an app. The device was built with a significant reduction in cost compared to qPCR systems. We anticipate that it will be most suitable in a professional setting such as a clinic.

Additionally, we took one step further by lyophilizing the one-pot reagent to facilitate transportation, distribution, and deployment without the need for cold-chain requirements in remote or austere settings. We obtained the lyo-ready Warmstart® reagent currently in development from New England Biolabs that is comprised of a glycerol-free LAMP reaction mixture. Prior to lyophilization, BrCas12b, sgRNA, and cryoprotectant were added to the lyo-ready mixture. Under our testing conditions, we observed a slightly reduced detection signal from BrCas12b to that of the glycerol version, and the LAMP exhibited a five-minute delay in reaction time (Fig. 2j,k), possibly due to the incorporation of several enzymes in the one-pot master mix.

## DISCUSSION

As the COVID-19 pandemic progresses with a growing number of VOCs circulating, the need for a rapid, one-pot detection system to differentiate among the strains is paramount^21-23^. Currently, the primary method to reliably detect VOCs is via next-generation sequencing (NGS)^24-26^. Although NGS-based methods are vital in the identification and confirmation of new variants, they are labor-intensive and require several hours of processing time, which limits their cost-effectiveness for real-time molecular epidemiology surveillance. Due to a number of advantages of BrCas12b such as high specificity, and robust trans-cleavage at RT-LAMP reaction temperatures, we applied this effector towards a one-pot detection reaction. BrCas12b was observed to have minimal inhibitory effects on RT-LAMP reaction and perform well in isothermal amplification buffer. In ≤ 30 minutes, samples with high viral load (C_t_ value ≤ 30) exhibited 100% accuracy, and we achieved ≥ 95% sensitivity detecting VOCs for samples with C_t_ value ≤ 32. When combined with UDG, the one-pot detection reaction minimizes carryover contamination as no false positives were observed in our clinical validation. Additionally, the freeze-drying process reduces the need for low-temperature logistics. We envision that CRISPR-SPADE with the portable detection instrument will move us closer to providing a cost-effective, rapid point-of-care test.

## MATERIALS AND METHODS

### Expression plasmid construction

BrCas12b gene sequence derived from *Brevibacillus* sp. SYP-B805 was obtained from the National Center for Biotechnology Information (GenBank ID: WP_165214399.1). The gene sequence was then codon-optimized using the GenSmart™ Codon Optimization tool (Genscript) for bacterial protein expression, synthesized by Twist Bioscience, and cloned into PET28a^+^ vector.

### Protein expression and purification

The BrCas12b expression plasmid was transformed into BL21(DE3) competent E. coli cells. Individual colonies were picked and inoculated in 8 mL Luria Broth (Fisher Scientific) for 12-15 hours. The culture was added to a 2 L of homemade Terrific Broth for scale-up and shaken until the OD600 reached 0.5 – 0.8. The culture was then placed on ice for 15 – 30 minutes prior to the addition of 0.2 mM IPTG (Isopropyl β-d-1-thiogalactopyranoside) followed by overnight expression at 16°C for 14-18 hours.

The next day, cells were harvested by centrifugation at 10,000 xg for 5 minutes. Cell pellets were then resuspended in lysis buffer (0.5 M NaCl, 50 mM Tris-HCl, pH = 7.5, 0.5 mM TCEP, 1mM PMSF, 0.25 mg/ml lysozyme, and 10 ug/mL Deoxyribose nuclease I). The suspended cell mixture was disrupted by sonication and centrifuged at 39,800 xg for 30 minutes at 4°C. The supernatant was collected and filtered through a 0.22 µm filter (Millipore Sigma) prior to injecting into a Histrap 5 mL FF column (Cytiva) pre-equilibrated with Wash Buffer (0.5 M NaCl, 50 mM Tris-HCl, pH = 7.5, 0.5 mM TCEP, 20 mM Imidazole). The purification process was performed via an FPLC Biologic Duoflow system (Bio-rad). After lysate injection and washing with Wash Buffer A, the column was eluted with 40 mL of Elution Buffer B(0.5 M NaCl, 50 mM Tris-HCl pH = 7.5, 0.5 mM TCEP, 300 mM Imidazole). The eluted fractions were pooled together, transferred to a 10 KDa - 14 KDa dialysis bag, and dialyzed in Dialysis Buffer (0.25 M NaCl, 40 mM HEPES, pH = 7, 1 mM DTT) overnight at 4°C.

Following overnight dialysis, the BrCas12b mixture was concentrated down to 15 mL in a 30 kDa MWCO Vivaspin® 20 concentrator via centrifugation with the speed of 2000 xg at 4°C. The concentrate was mixed at 1:1 ratio with Buffer C (150 mM NaCl, 50 mM HEPES, pH = 7, 0.5 mM TCEP) before injecting into a Hitrap Heparin 1 mL HP column (Cytiva). The protein was eluted over a gradient buffer exchange from Buffer C to Buffer D (2 M NaCl, 50 mM HEPES, pH = 7, 0.5 mM TCEP). Purest elution fractions of BrCas12b were collected, concentrated down, snapped frozen, and stored at -80°C until use. Experiment-ready BrCas12b was diluted in storage buffer (500 mM NaCl, 20 mM sodium acetate, 0.1 mM EDTA, 0.1 mM TCEP, 50% Glycerol, pH 7 @ 25°C) which can be stored at -20°C.

AacCas12b and AapCas12b expression plasmids were obtained as a gift from Jennifer Doudna (Addgene plasmid # 113433) and Wei Li (Addgene plasmid # 121949), respectively. These two proteins were expressed and purified following Chen *et al*^*8*^. and Teng *et al*^*27*^.

### Nucleic acid preparation

For in vitro cleavage assay and differential scanning fluorimetry experiments, single-guide RNA (sgRNA) was synthesized using PCR (Takara) and in vitro transcription (IVT) using the HiScribe™ T7 Quick High Yield RNA Synthesis Kit (New England Biolabs). The IVT reaction was purified using the RNA Clean and Concentrator Kit (Zymo Research).

For patient sample detection, the sgRNA was either chemically synthesized by Integrated DNA Technologies (IDT) or enzymatically synthesized by an in vitro transcription (IVT) reaction followed by HPLC purification via the HPLC 1100 system (Agilent). The control genomic RNAs for Beta and Delta variants were obtained from Twist Bioscience. The control genomic RNA for wild-type SARS-CoV-2 and Alpha were obtained from BEI Resources. Control genomic RNA for Gamma was obtained from Salemi Lab at the University of Florida.

### *In vitro* dsDNA cleavage assay

Cas12b, sgRNA, and dsDNA were combined in 1x NEBuffer 2.1 on ice to a final concentration of 100 nM, 125 nM, and 7nM, respectively. The reaction mixture was immediately transferred to a pre-set thermocycler and incubated for 1 hour at different temperatures (37°C, 47°C, 52°C, 57°C, 59°C, 61°C, 63°C, 65°C, 70°C, and 75°C). After incubation, the reaction mixture was treated with a 6x quenching buffer (30% glycerol, 1.2% SDS, 250 mM EDTA). The reaction products were analyzed on 1% agarose gel pre-stained with GelRed (Biotium).

### Differential scanning fluorimetry

The reaction was performed following Nguyen *et al*^*28*^. In short, Cas12b and sgRNA were combined in a 1:2.5 ratio (1 µM: 2.5 µM final concentration) in a mixture of Protein Thermal Shift™ buffer (Thermofisher) and 1x reaction buffer (100 mM NaCl, 50 nM Tris-HCl, pH = 7.5, 1 mM DTT, and 10 mM MgCl_2_). The reaction was incubated at 37°C for 30 minutes to ensure the complexation of sgRNA and Cas12b prior to adding the Protein Thermal Shift™ dye (Thermofisher). The reaction mixture was then transferred to the qPCR StepOne Plus system (Thermofisher), and the binary complex temperature melting profile was recorded over a temperature range of 25-99°C with a ramp rate of 1%/second. The experiment was carried in duplicates and repeated twice.

### Cas12b trans-cleavage kinetic assay

The trans-cleavage kinetic experiment was carried out following Cofsky *et al*^*29*^. with modifications. In short, BrCas12b, sgRNA, and dsDNA activator were combined to a final concentration of 100 nM: 125 nM: 1 nM respectively in 1x NEBuffer 2.1 (New England Biolabs) and incubated at 62°C for 30 minutes. HEX-polyT-Quencher reporter (FQ) at various concentrations (10 nM, 100 nM, 200 nM, 500 nM, 1 µM, and 2 µM) was added to the reaction mixture containing Cas12b trans activated complex, and the entire reaction was immediately transferred to a plate reader. Fluorescence measurements (λ_ex_: 483/30 nm, λ_em_: 530/30 nm) were read every 30 seconds using the Biotek Synergy Neo (Agilent) that was pre-heated to 62°C. Initial velocity for each FQ concentration was determined by establishing the slope for all components and subtracting the slope for the no-activator control. The cleaved HEX-polyT reporter was titrated to different concentrations (10 nM, 100 nM, 200 nM, 500 nM, 1 µM, and 2 µM) and measured in the same experiment for the conversion of FQ fluorescence to concentration, eliminating non-linearity at high reporter concentrations.

### Patient sample collection

Saliva samples were collected from the University of Florida via Marco Salemi Lab in the Department of Pathology, Immunology and Laboratory Medicine, College of Medicine. The sample collection followed the guidelines approved by UF Institutional Review Board (IRB202000781) and through Evaluating the Molecular Epidemiology of Coronavirus (COVID-19) in Florida (IRB202000633). Samples were extracted following CDC-recommended procedures and sequenced (described below) to identify SARS-CoV-2 lineages.

### RNA extraction, library preparation, and sequencing

Viral RNA was extracted from 180 µl of each saliva sample using the QIAamp 96 Viral RNA Kit with the QIAcube HT (Qiagen) using the following settings with a filter plate: the lysed sample was pre-mixed 8 times before subjecting to vacuum for 5 minutes at 25 kPa and vacuum for 3 min at 70 kPa. Following 3 washes using the same vacuum conditions above, the samples were eluted in 75 µl AVE buffer followed by a final vacuum for 6 minutes at 60 kPa. Next, Nine microliters of RNA were used for cDNA synthesis and library preparation using the Illumina COVIDSeq Test kit (Illumina) and Mosquito HV Genomics Liquid Handler (SPT Labtech Inc.). The size and purity of the library were determined using the 4200 TapeStation System (Agilent) and the Qubit dsDNA HS Assay Kit (Life Technologies) according to the manufacturer’s instructions. Constructed libraries were pooled and sequenced using the NovaSeq 6000 Sequencing System SP Reagent Kit and the NovaSeq Xp 2-Lane Kit. Illumina’s DRAGEN pipeline was used to derive sample consensus sequences, which were filtered based on a minimum of 70% coverage of the genome.

### SARS-CoV-2 viral load quantification

Levels of SARS-CoV-2 were determined using the 2019-nCoV_N1 assay (primer and probe set) with 2019-nCoV_N_positive control (IDT). Viral RNA was extracted as previously described then subjected to first-strand synthesis using ProtoScript II Reverse Transcriptase according to the manufacturer’s instructions (New England Biolabs). Quantitative PCR was performed using TaqMan Fast Advanced Master Mix (Waltham) according to the manufacturer’s instructions. A standard curve was generated using N1 quantitative standards 10-fold diluted to determine viral copies. The assay was run in triplicate including one non-template control.

### One-pot BrCas12b detection assay

LAMP primers targeting wild-type SARS-CoV-2 and variants of concern were designed using NEB® LAMP Primer Design Tool (New England Biolabs) and PrimerExplorer v5 at https://primerexplorer.jp/e/^14^.

For the one-pot detection assay that tracks both target amplification and BrCas12b trans-cleavage, the reaction was assembled by combining the following reagents:

| Item # | Reagents | Volume (μL) | Final Concentration |
| --- | --- | --- | --- |
| 1 | WarmStart® Multi-Purpose LAMP/RT-LAMP 2X Master Mix (with UDG) | 12.5 | 1X |
| 2 | 10X LAMP Primer Mix | 2.5 | 1X |
| 3 | 100 μM HEX-polyT-Quencher Reporter | 0.5 | 2 μM |
| 4 | 25 μM SYTO™ 9 Dye | 1 | 1 μM |
| 5 | 10 μM BrCas12b | 0.25 | 0.1 μM |
| 6 | 10 μM sgRNA | 0.5 | 0.2 μM |
| 7 | RNase-free Water | 2.75 | ----- |
|  | **Total** | **20** |  |

For the rapid one-pot detection assay monitoring BrCas12b trans-cleavage only, the reaction was assembled by combining the following reagents:

| Item # | Reagents | Volume (μL) | Final Concentration |
| --- | --- | --- | --- |
| 1 | WarmStart® Multi-Purpose LAMP/RT-LAMP 2X Master Mix (with UDG) | 12.5 | 1X |
| 2 | 10X LAMP Primer Mix | 2.5 | 1X |
| 3 | 100 μM HEX-polyT-Quencher Reporter | 0.5 | 2 μM |
| 4 | 10 μM BrCas12b | 0.25 | 0.1 μM |
| 5 | 10 μM sgRNA | 0.5 | 0.2 μM |
| 6 | RNase-free Water | 3.75 | ----- |
|  | **Total** | **20** |  |

For SARS-CoV-2 variant specificity testing, the one-pot reaction was added with LAMP primer and sgRNA corresponding to a SARS-CoV-2 variant and tested against other variants. For Limit of Detection (LoD) testing, RNA controls were subjected to serial dilution in triplicates and added to the one-pot reaction. For patient sample detection testing, 5 µL of extracted nucleic acid from clinical samples were added to the one-pot reaction.

The reaction mixture was transferred to a Bio-rad CFX96 Real-Time system with a C1000 Thermal Cycler module. The reaction was isothermally incubated at 62°C. Fluorescence measurements were read every 30 seconds per cycle over 90 cycles. The FAM channel (λex: 470/20 nm, λem: 520/10 nm) was used to monitor SYTO9™, and the HEX channel (λex: 525/10 nm, λem: 570/10 nm) was used to detect BrCas12b trans-cleavage via FRET-based HEX reporter.

### Lyophilization of BrCas12a one-pot detection assay

Developmental lyo-ready Warmstart® Master Mix with Uracil-DNA glycosylases (UDG) (New England Biolabs) was combined with 100 nM BrCas12b, 200 nM sgRNA, and 1.25 µmoles Trehalose in ice. The mixture was placed in an aluminum cooling block and kept in dry ice for 30 minutes. The cooling block containing samples was then transferred to a freeze-drying system (Labconco) and lyophilized for 4 hours. To initiate the reaction, the lyophilized master mix was reconstituted in 20 µL RNase-free water.

### Imaging using a mobile phone and a lens

One-pot BrCas12b reagents were combined in an optically clear PCR tube (Applied Biosystems) followed by the addition of the extracted RNA sample and 62°C incubation for 20 minutes. The reaction tube was then imaged using a battery-operated 410 nm – 415 nm UVA-blue flashlight on a mobile phone in a dark setting. To visualize a sample containing FRET-based HEX reporter, the imaging system was assembled by attaching a combination of yellow and orange lenses onto the flashlight camera (NestEcho).

### Portable multiplexing detection prototype (FISSH)

To build a portable detection device that is capable of multiplexing (in this study the device enables monitoring both LAMP amplification and BrCas12b collateral cleavage), the optical system was assembled with two wavelengths. The first one was an MCPCB-mounted LED on 490 nm with a bandwidth of 26 nm, 205 mW driven by 199 mA with impulse length 150 ms for exposure of FITC (Thorlabs). The second LED worked on 625 nm with a bandwidth of 17 nm, 700 mW driven by 215 mA with the same length of impulse for exposure of Cy5 (Thorlabs). The beam of light from the LED was collimated (straightened) by the lens before it traveled to the dichroic mirror and reflected the light into an excitation filter, which cut off the parasitic light. The light beam went to the second dichroic mirror which reflected the beam of light to the collimating lens, concentrating light into the vial with the sample. When the sample started emitting the light from the fluorophore, it then traveled through the collimating lens into the second dichroic mirror, allowing the beam light to go through directly into the emission filter, cutting off the parasitic light. Next, the beam went through another collimating lens prior to being focused and detected by a photodiode sensor/detector (Thorlabs). A schematic diagram of the device is presented in figure 2e, and the specifications of device parts are indicated in supplementary table S1.

## Data Availability

The BrCas12b expression plasmid has been deposited and will be made available on Addgene (plasmid #170819). Sequencing data of tested patient samples have been deposited into GISAID and will be available in the near future.

## AUTHOR CONTRIBUTIONS

P.K.J and L.T.N designed the experiments. L.T.N, N.C.M, B.L.M.P carried out experiments and performed data analyses. L.T.N, B.L.M.P, and N.C.M wrote the primary manuscript. M.S. and R.D. provided the patient samples. M.N.C. performed the sequencing analysis of patient samples. J.K. and J.S. co-developed the portable multiplexing device (FISSH). The manuscript was edited, refined, and approved by all authors.

## ACKNOWLEDGMENTS

We are thankful to Dr. Christopher Dervinis in the Forest Genomics group and Dr. Whitney Stoppel in the department of Chemical Engineering for the use of the lyophilizers. Additionally, we appreciate the support from New England Biolabs for the use of lyo-ready Warmstart® reagents. Finally, we are grateful to the Health Care Center at the University of Florida and members of Jain Lab for their insightful feedback. The following reagents were obtained through BEI Resources, NIAID, NIH: Genomic RNA from SARS-Related Coronavirus 2, Isolate USA-WA1/ 2020, NR-52285, (Lineage B.1.1.7), NR-55244, contributed by the Centers for Disease Control and Prevention. This work was supported in part by funds from the University of Florida Office of Research and Health Science Center with resources from the Interdisciplinary Center for Biotechnology Research Gene Expression Core (RRID: SCR_019145), NextGen Sequencing Core (RRID: SCR_019152), and Bioinformatics Core (RRID: SCR_019120).

## DATA AVAILABILITY

The BrCas12b expression plasmid has been deposited and will be made available on Addgene (plasmid #170819). Sequencing Data of tested patient samples have been deposited into GISAID.

