## Supplementary Information for "A Thermostable Cas12b from *Brevibacillus* Leverages One-pot Detection of SARS-CoV-2 Variants of Concern"

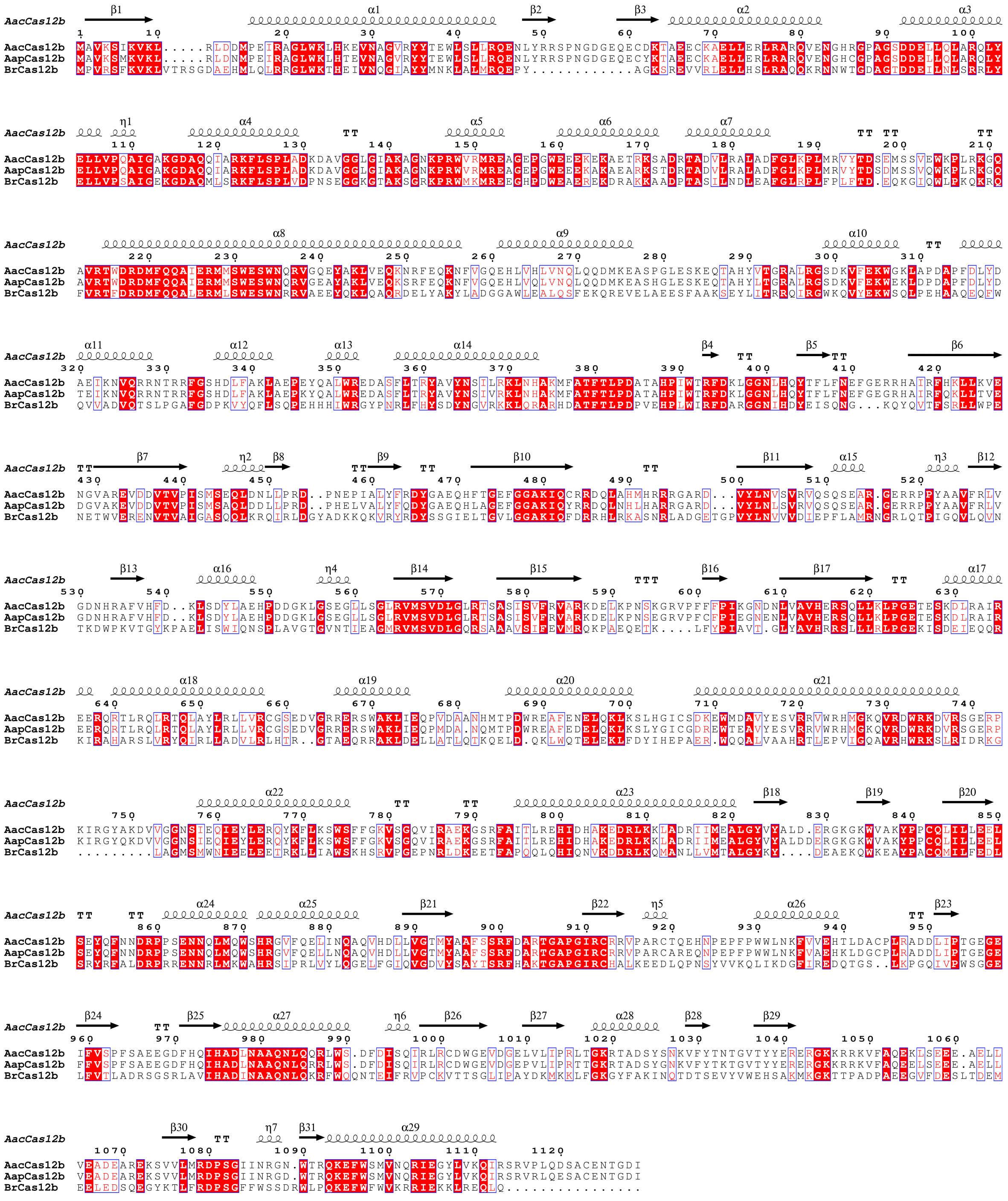

**Figure S1**. **Sequence alignment of AapCas12 and BrCas12b against the structure of AacCas12b.** The sequences of the above effectors were aligned using MultiAlin with Blosum62-12-2. The aligned sequence file was imported into Espript 3.0[30] and aligned against the AacCas12b structure (PDBID: 5U30)[31].

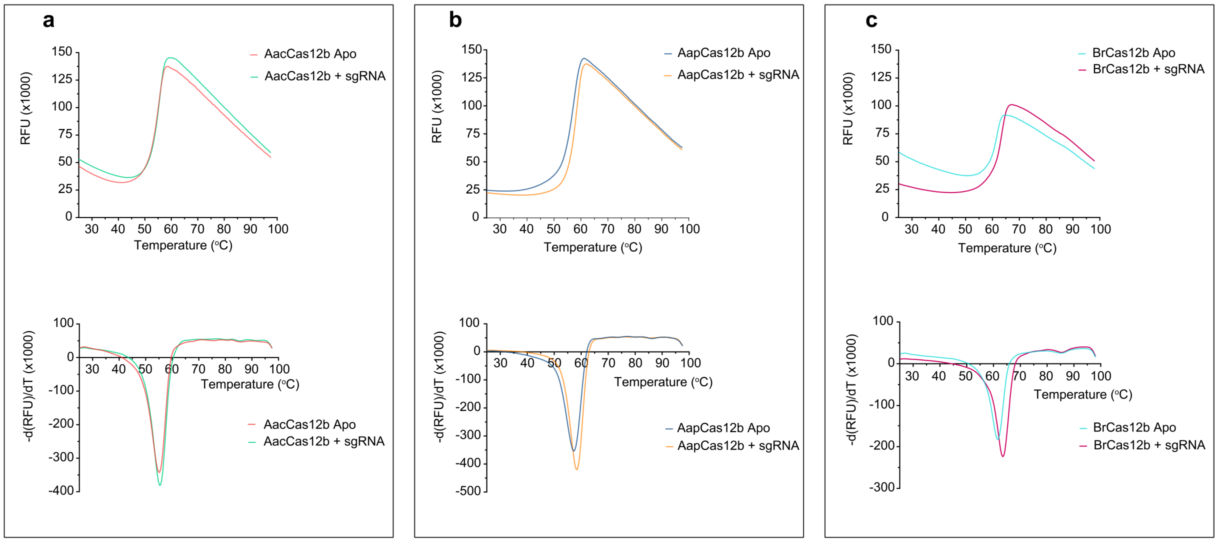


**Figure S2**. **Differential scanning fluorimetry of Apo and binary complexes of AacCas12b, AapCas12b, and BrCas12b**. The reaction was performed in duplicates and repeated twice (n = 2). The melting temperature was determined based on the global minimum of derivative fluorescence with respect to temperature.

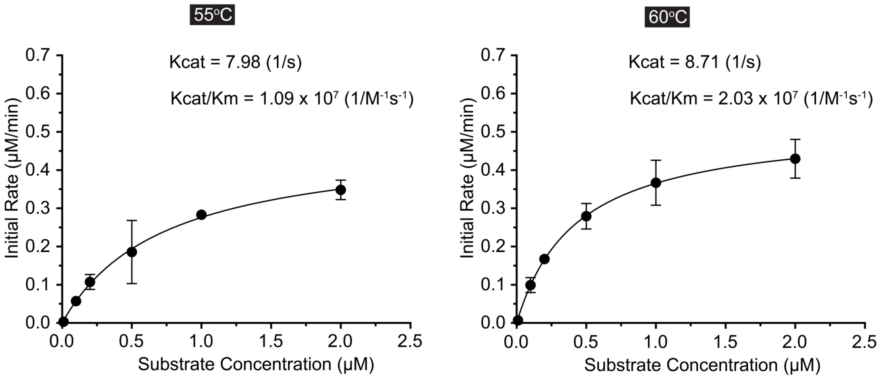


**Figure S3**. **Trans-cleavage kinetic analysis of BrCas12b.** (**a**) at 55ºC and (**b**) at 60ºC, (n = 2).

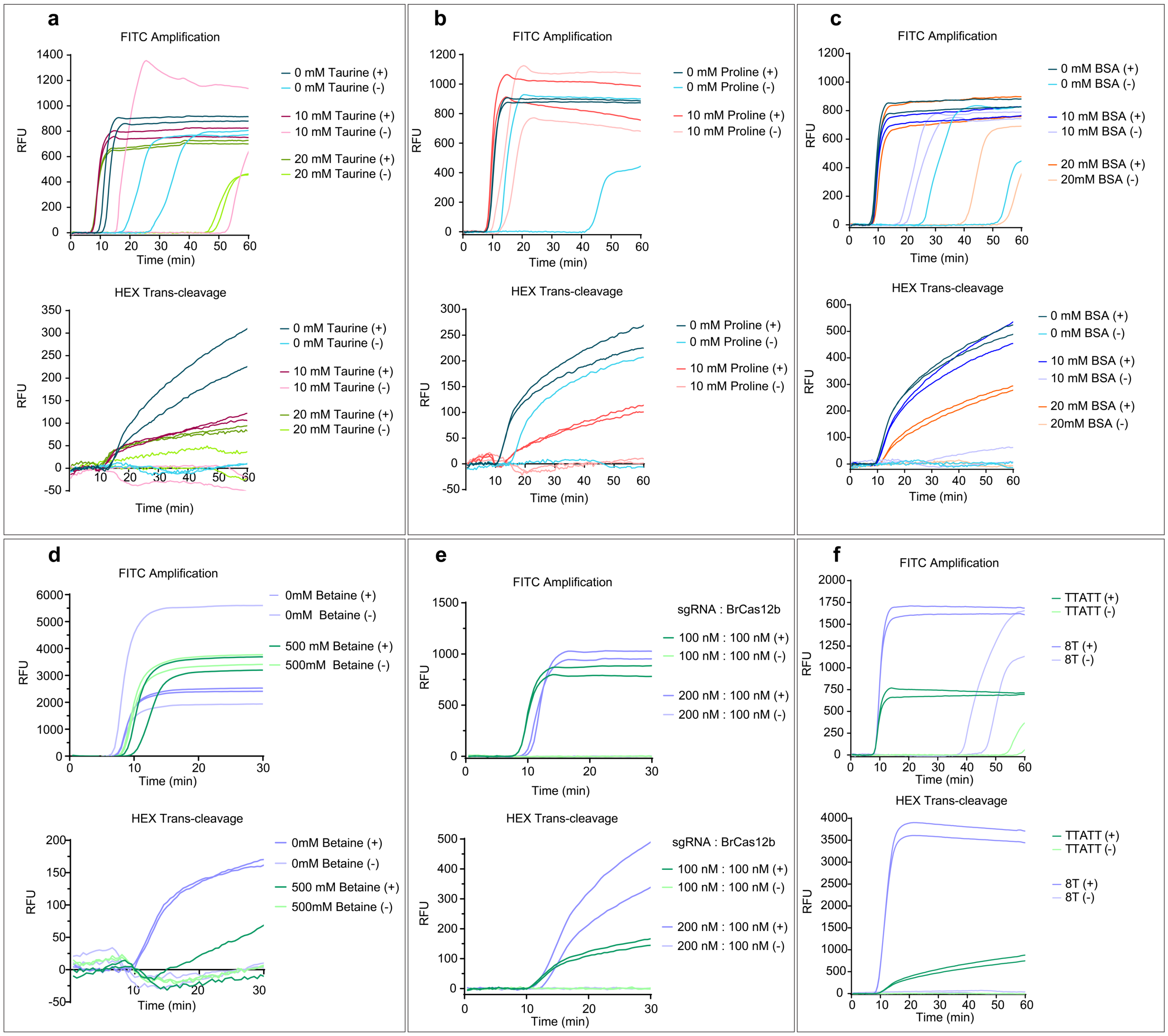


**Figure S4**. **BrCas12b one-pot reaction optimization**. (**a**) – (**f**) Various additives including taurine, proline, bovine serum albumin (BSA), and betaine were used in (a), (b), (c), and (d). Additionally, sgRNA, BrCas12b concentrations, and reporters were also optimized in (e) and (f). FAM channel indicates LAMP amplification detecting SYTO$^{TM}$ 9, and HEX channel was used to detect BrCas12b trans-cleavage activity.

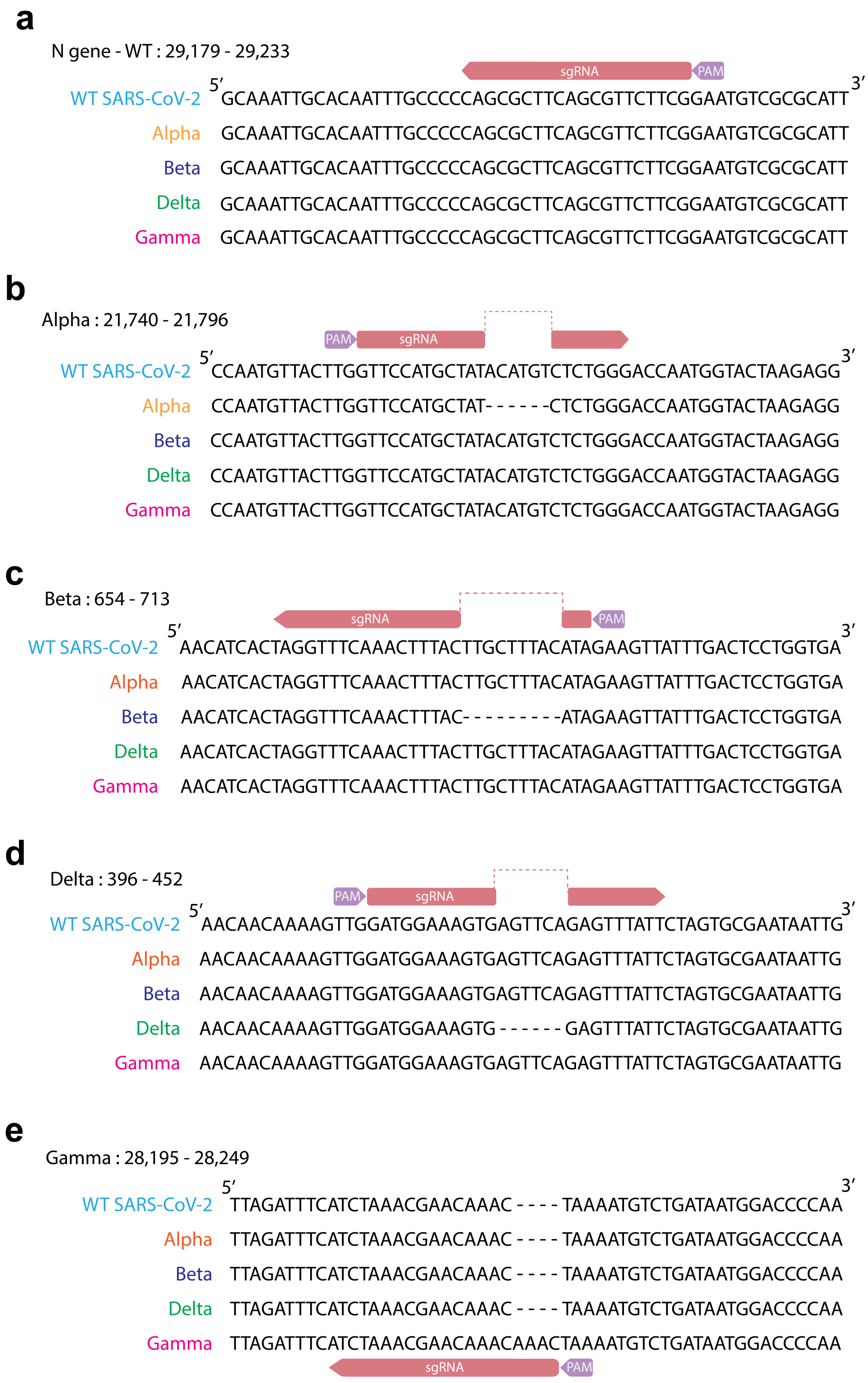


**Figure S5**. Specificity of single-guide RNA designs among the tested SARS-CoV-2 variants of concerns.

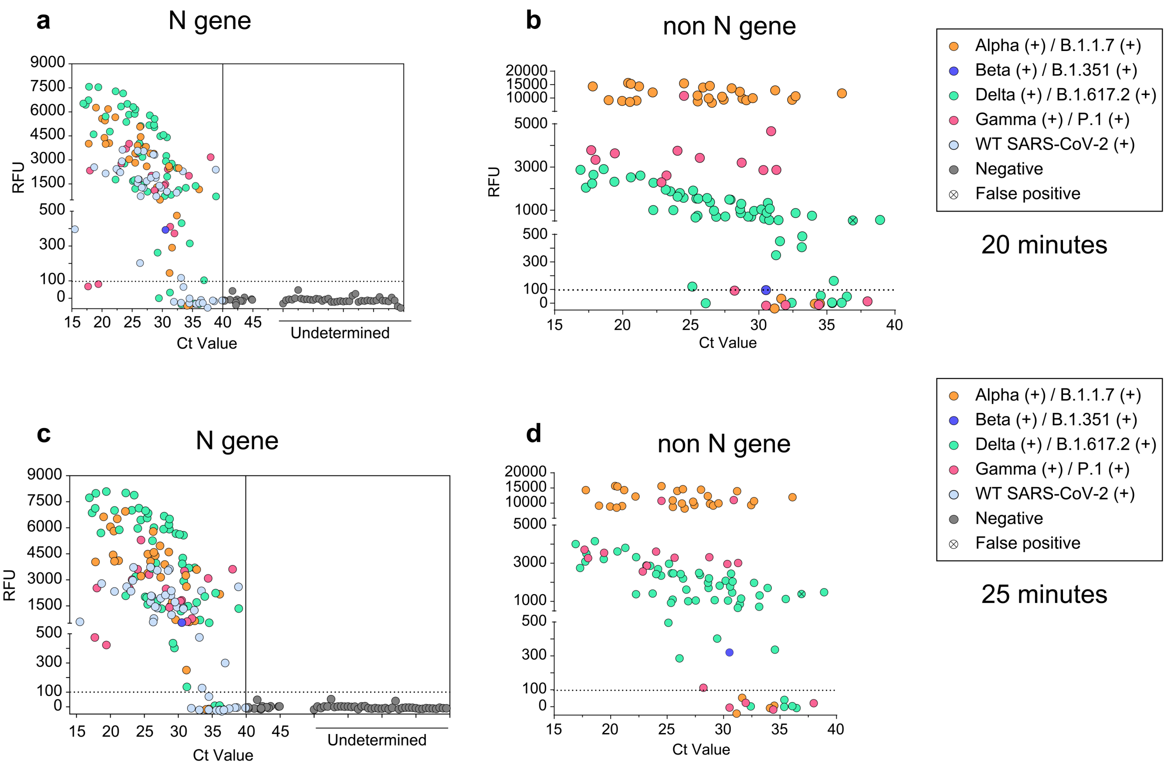


**Figure S6**. **Clinical validation of one-pot BrCa12b detection assay (supplement to figure 2 in the main text).** (**a**) and (**b**) One-pot patient sample detection with fluorescence measurements taken at t = 20 minutes. (**c**) and (**d**) The same assay as in (a) and (b) but at t = 25 minutes. The N gene target indicates the presence of SARS-CoV-2 while non-N gene targets indicates detection of variants.

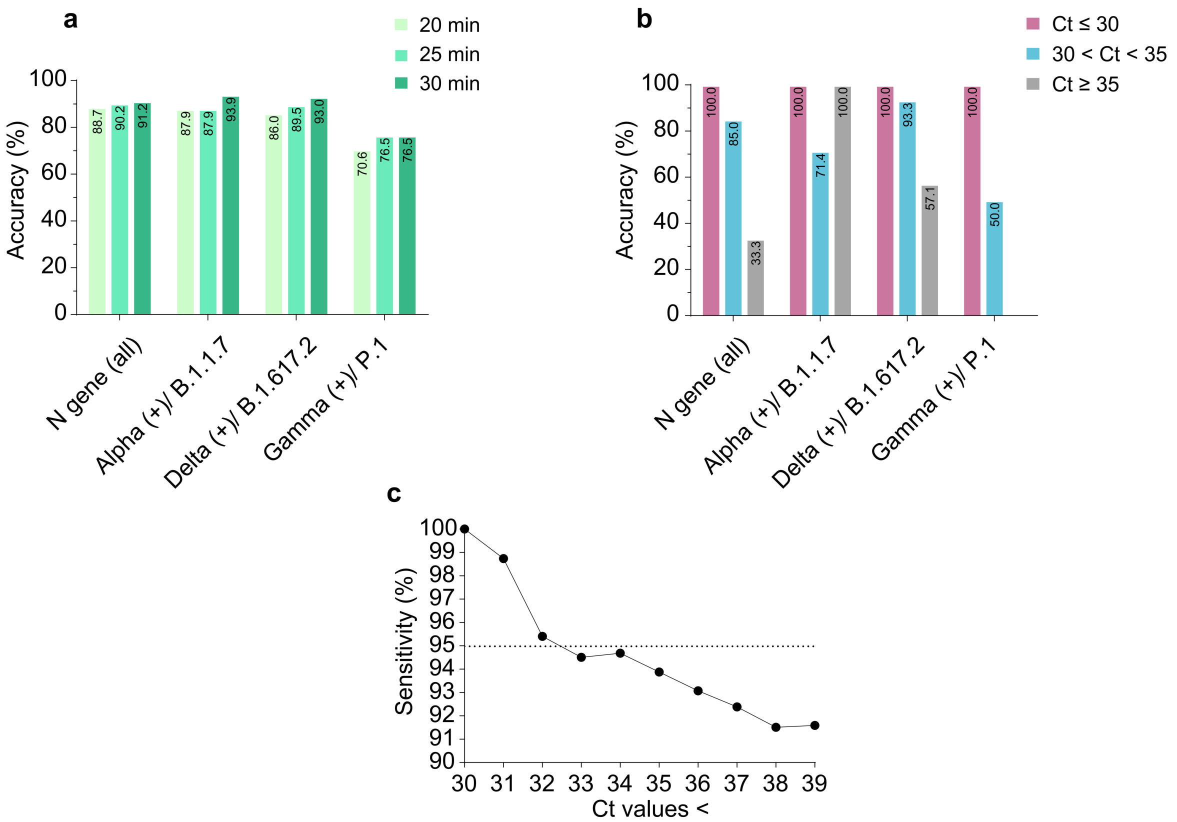


**Figure S7**. **Percent accuracy of patient sample validation.** (**a**) Percent accuracy of variant detection with respect to time. (**b**) Percent accuracy of variant detection with respect to Ct values. (**c**) Cumulative sensitivity of VOC patient sample detection with respect to $C_t$ values. Calculations were performed at t = 30 minutes.

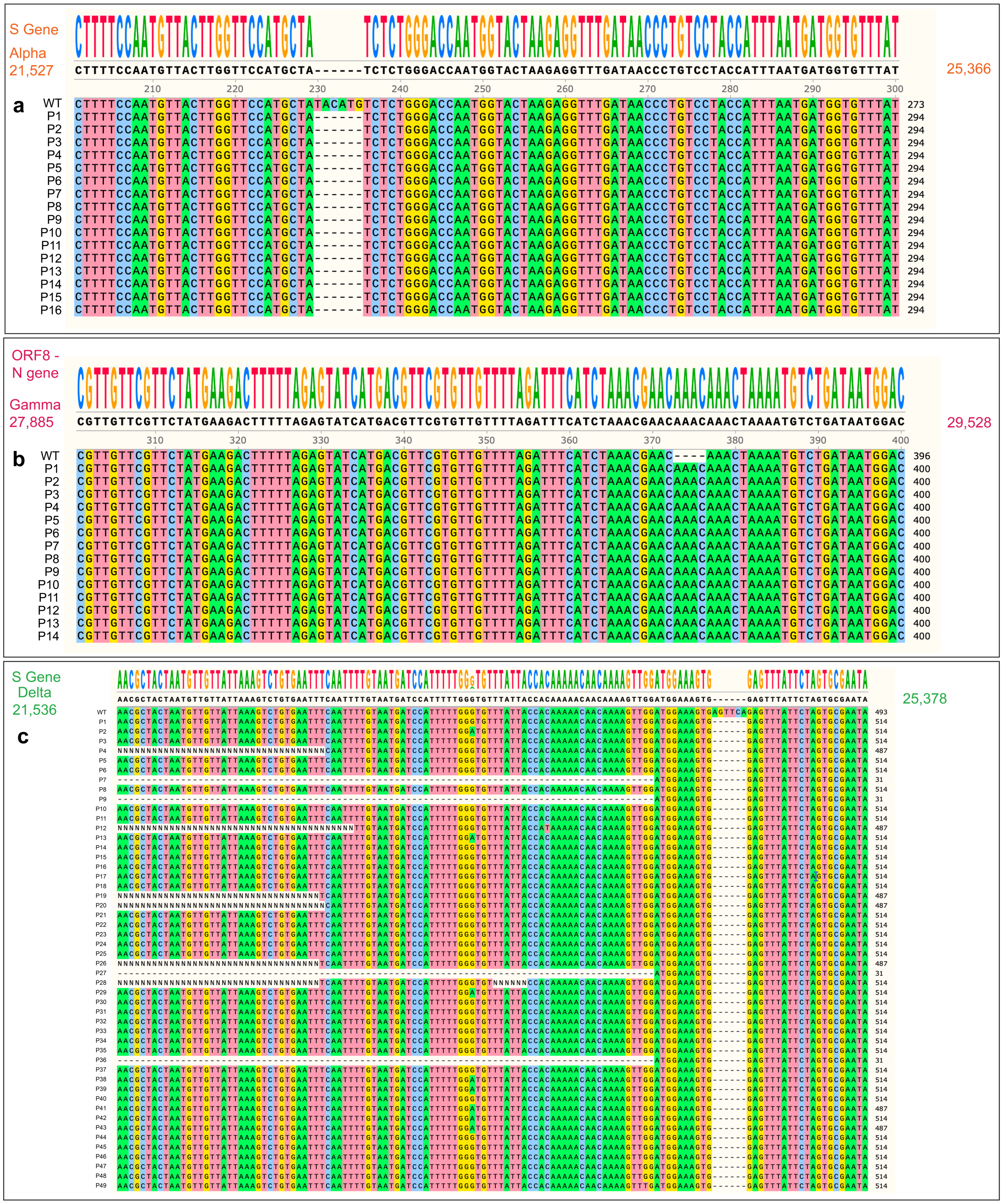


**Figure S8**. **Sequence alignment of patient samples in reference to wild-type SARS-CoV-2 genomic RNA.** Samples were confirmed through sequencing to identify SARS-CoV-2 variants. (**a**) Alpha. (**b**) Gamma. (**c**) Delta. “P” represents a sample. Sequences were imported into Snapgene and aligned with Clustal Omega[32,33].

**Table S1. Portable multiplexing detection device (FISSH) specifications**

**Dichroic Beam Splitter (Mirror) 1 (close to both LEDs on the scheme)**

| Specification | Value |
|---|---|
| Reflection Band | $R_{avg}$ > 98% 350 – 585 nm |
| Transmission Band | $T_{avg}$ > 93% 601 – 950 nm |

**Dichroic Beam Splitter (Mirror) 1 (close to sensor and sample)**

| | |
|---|---|
| Reflection Band 1 | $R_{abs}$ > 94% 471 – 491 nm |
| Reflection Band 2 | $R_{abs}$ > 94% 541.5 – 544.5 nm |
| Reflection Band 3 | $R_{abs}$ > 94% 632.8 – 647.1 nm |
| Transmission Band 1 | $T_{avg}$ > 93% 503.5 – 526.5 nm |
| Transmission Band 2 | $T_{avg}$ > 93% 560 – 615.5 nm |
| Transmission Band 3 | $T_{avg}$ > 93% 665.5 – 800 nm |

**Excitation Filter**

| | |
|---|---|
| Transmission Band 1 | $T_{avg}$ > 85% 352 – 404 nm |
| Center Wavelength 1 | 377.5 nm |
| Guaranteed Minimum Bandwidth 1 | 52 nm |
| FWHM Bandwidth 1 (nominal) | 57.3 nm |
| Transmission Band 2 | $T_{avg}$ > 93% 461 – 487.5 nm |
| Center Wavelength 2 | 474.1 nm |
| Guaranteed Minimum Bandwidth 2 | 26.5 nm |
| FWHM Bandwidth 2 (nominal) | 30.1 nm |
| Transmission Band 3 | $T_{avg}$ > 93% 543 – 566 nm |
| Center Wavelength 3 | 554.5 nm |
| Guaranteed Minimum Bandwidth 3 | 23 nm |
| FWHM Bandwidth 3 (nominal) | 27.6 nm |
| Transmission Band 4 | $T_{avg}$ > 93% 626 – 644 nm |
| Center Wavelength 4 | 635 nm |
| Guaranteed Minimum Bandwidth 4 | 18 nm |
| FWHM Bandwidth 4 (nominal) | 22.9 nm |

**Emission Filter**

| | |
|---|---|
| Transmission Band 1 | $T_{avg}$ > 93% 414 – 450 nm |

| | |
|---|---|
| Center Wavelength 1 | 432 nm |
| Guaranteed Minimum Bandwidth 1 | 36 nm |
| FWHM Bandwidth 1 (nominal) | 39.7 nm |
| Transmission Band 2 | $T_{avg}$ > 93% 499.5 – 530 nm |
| Center Wavelength 2 | 514.8 nm |
| Guaranteed Minimum Bandwidth 2 | 30.5 nm |
| FWHM Bandwidth 2 (nominal) | 35.5 nm |
| Transmission Band 3 | $T_{avg}$ > 93% 580 – 611 nm |
| Center Wavelength 3 | 595.5 nm |
| Guaranteed Minimum Bandwidth 3 | 31 nm |
| FWHM Bandwidth 3 (nominal) | 36.2 nm |
| Transmission Band 4 | $T_{avg}$ > 93% 661 – 800 nm |
| Center Wavelength 4 | 730.5 nm |
| Guaranteed Minimum Bandwidth 4 | 139 nm |
| FWHM Bandwidth 4 (nominal) | 147.2 nm |

**Table S2. Sequences used in the study**

**A. RT-LAMP Primers**

| Primer target | Primer name | Sequence | 1X Concentration |
|---|---|---|---|
| N gene (for presence of SARS-CoV-2) | N_F3 | AACACAAGCTTTCGGCAG | 0.2 μM |
| | N_B3 | GAAATTTGGATCTTTGTCATCC | 0.2 μM |
| | N_FIP | TGCGGCCAATGTTTGTAATCAGCC AAGGAA ATTTTGGGGAC | 1.6 μM |
| | N_BIP | CGCATTGGCATGGAAGTCACTTTG ATGGC ACCTGTGTAG | 1.6 μM |
| | N_LF | TTCCTTGTCTGATTAGTTC | 0.8 μM |
| | N_LB | ACCTTCGGGAACGTGGTT | 0.8 μM |
| Alpha (B.1.1.7) | Alpha_F3 | ATACACTAATTCTTTCACACGT | 0.2 μM |
| | Alpha_B3 | CCTCTTATTATGTTAGACTTCTCAG | 0.2 μM |
| | Alpha_FIP | GGAAAAGAAAGGTAAGAACAACCC TGACAAAGTTTTCAGAT | 1.6 μM |
| | Alpha_BIP | ATGGTACTAAGAGGTTTGATTGGA AGCAAAATAAACACCATC | 1.6 μM |
| | Alpha_LF | CCTGAGTTGAATGTAAAACTGAGG | 0.8 μM |
| | Alpha_LB | AACCCTGTCCTACCATTTAA | 0.8 μM |
| Beta (B.1.351) | Beta_F3 | ACACGCCTATTAATTTAGTGC | 0.4 μM |
| | Beta_B3 | TAGAAAAGTCCTAGGTTGAAGA | 0.4 μM |
| | Beta_FIP | CCTAGTGATGTTAATACCTATTGGC TCCCTCAGGGTTTTTCG | 1.6 μM |

| | | | |
|---|---|---|---|
| | Beta_BIP | AAGTTATTTGACTCCTGGTCATAAT AAGCTGCAGCACC | 1.6 μM |
| | Beta_LF | CAAATCTACCAATGGTTCTAAAGC | 0.8 μM |
| | Beta_LB | GATTCTTCTTCAGGTTGGACAGC | 0.8 μM |
| Delta (B.1.617.2) | Delta_F3 | CCCTACTTATTGTTAATAACGCT | 0.4 μM |
| | Delta_B3 | ATTCTTAAACACAAATTCCCTAAG | 0.4 μM |
| | Delta_FIP | TTTTTGTGGTAATAAACACCCAAAA ACTAATGTTGTTATTAAAGTCTGTG | 1.6 μM |
| | Delta_BIP | CTAGTGCGAATAATTGCACTTTTGA TGTTTTCCTTCAAGGTCCAT | 1.6 μM |
| | Delta_LF | ATGGATCATTACAAAATTGAAA | 0.8 μM |
| | Delta_LB | ATATGTCTCTCAGCCTTTTCTT | 0.8 μM |
| Gamma (P.1) | Gamma_F3 | CAATTAATTGCCAGGAACCTAA | 0.2 μM |
| | Gamma_B3 | TACTGCCAGTTGAATCTGA | 0.2 μM |
| | Gamma_FIP | CAACACGAACGTCATGATACTCTA AGGGTAGTCTTGTAGTG | 1.6 μM |
| | Gamma_BIP | ACTAAAATGTCTGATAATGGACCC CGGGTCCACCAAACGTAAT | 1.6 μM |
| | Gamma_LF | AAAGTCTTCATAGAACGAACAA | 0.8 μM |
| | Gamma_LB | AAAATCAGCGAAATGCACCCC | 0.8 μM |
| RNase P | RNaseP_F3 | TTGATGAGCTGGAGCCA | 0.2 μM |
| | RNaseP_B3 | CACCCTCAATGCAGAGTC | 0.2 μM |
| | RNaseP_FIP | GTGTGACCCTGAAGACTCGGTTTT AGCCA CTGACTCGGATC | 1.6 μM |
| | RNaseP_BIP | CCTCCGTGATATGGCTCTTCGTTTT TTTCTT ACATGGCTCTGGTC | 1.6 μM |
| | RNaseP_LF | ATGTGGATGGCTGAGTTGTT | 0.8 μM |
| | RNaseP_LB | CATGCTGAGTACTGGACCTC | 0.8 μM |

**B. Single-guide RNA**

| Target | Sequence |
|---|---|
| N gene (for presence of SARS-CoV-2) | GAAGGUGGUUAGCUACAGGCUGACCAGUGCAGUUGUGUCAUGUGCUACGGUGACCUAACACGUCACUCAGUCACAACGGCUAUCUAUAUUUCCACUAACCAAAGUUAGUGGAAAUGUAGAUGGUUAGCAC**<u>CGAAGAACGCUGAAGCGCUG</u>** |
| Alpha (B.1.1.7) | GAAGGUGGUUAGCUACAGGCUGACCAGUGCAGUUGUGUCAUGUGCUACGGUGACCUAACACGUCACUCAGUCACAACGGCUAUCUAUAUUUCCACUAACCAAAGUUAGUGGAAAUGUAGAUGGUUAGCAC**<u>GUUCCAUGCUAUCUCUGGGA</u>** |
| Beta (B.1.352) | GAAGGUGGUUAGCUACAGGCUGACCAGUGCAGUUGUGUCAUGUGCUACGGUGACCUAACACGUCACUCAGUC |

| | |
|---|---|
| | ACAACGGCUAUCUAUAUUCCACUAACCAAAGUUAGU GGAAAUGUAGAUGGUUAGCAC**UAUGUAAAGUUUGAA ACCUA** |
| Delta (B.1.617.2) | GAAGGUGGUUAGCUACAGGCUGACCAGUGCAGUUGU GUCAUGUGCUACGGUGACCUAACACGUCACUCAGUC ACAACGGCUAUCUAUAUUCCACUAACCAAAGUUAGU GGAAAUGUAGAUGGUUAGCAC**GAUGGAAAGUGGAGU UUAUU** |
| Gamma (P1) | GAAGGUGGUUAGCUACAGGCUGACCAGUGCAGUUGU GUCAUGUGCUACGGUGACCUAACACGUCACUCAGUC ACAACGGCUAUCUAUAUUCCACUAACCAAAGUUAGU GGAAAUGUAGAUGGUUAGCAC**GUUUGUUUGUUCGUU UAGAU** |
| RNase P | GAAGGUGGUUAGCUACAGGCUGACCAGUGCAGUUGU GUCAUGUGCUACGGUGACCUAACACGUCACUCAGUC ACAACGGCUAUCUAUAUUCCACUAACCAAAGUUAGU GGAAAUGUAGAUGGUUAGCAC**AAUUACUUGGGUGUG ACCCU** |

**C. Reporters**

| **Reporter name** | **Sequence** |
|---|---|
| Reporter 1 | /5HEX/TTTTTTTT/3IABkFQ/ |
| Reporter 2 | /5HEX/TTTTTTTT |
| Reporter 3 | /56FAM/TTATT/3IABkFQ/ |
| Reporter 4 | /56FAM/TTTTTTTT/3IABkFQ/ |
| Reporter 5 | 5Cy5/TTTTTTTT/3BHQ_2/ |

**D. Protein Sequences**

| **Name** | **Sequence** |
|---|---|
| AacCas12b | MAVKSIKVKLRLDDMPEIRAGLWKLHKEVNAGVRYYTEWLSLLRQE NLYRRSPNGDGEQECDKTAEECKAELLERLRARQVENGHRGPAG SDDELLQLARQLYELLVPQAIGAKGDAQQIARKFLSPLADKDAVGGL GIAKAGNKPRWVRMREAGEPGWEEEKEKAETRKSADRTADVLRAL ADFGLKPLMRVYTDSEMSSVEWKPLRKGQAVRTWDRDMFQQAIE RMMSWESWNQRVGQEYAKLVEQKNRFEQKNFVGQEHLVHLVNQ LQQDMKEASPGLESKEQTAHYVTGRALRGSDKVFEKWGKLAPDAP FDLYDAEIKNVQRRNTRRFGSHDLFAKLAEPEYQALWREDASFLTR YAVYNSILRKLNHAKMFATFTLPDATAHPIWTRFDKLGGNLHQYTFL FNEFGERRHAIRFHKLLKVENGVAREVDDVTVPISMSEQLDNLLPR |

| | |
|---|---|
| | DPNEPIALYFRDYGAEQHFTGEFGGAKIQCRRDQLAHMHRRRGAR DVYLNVSVRVQSQSEARGERRPPYAAVFRLVGDNHRAFVHFDKLS DYLAEHPDDGKLGSEGLLSGLRVMSVDLGLRTSASISVFRVARKDE LKPNSKGRVPFFFPIKGNDNLVAVHERSQLLKLPGETESKDLRAIRE ERQRTLRQLRTQLAYLRLLVRCGSEDVGRRERSWAKLIEQPVDAA NHMTPDWREAFENELQKLKSLHGICSDKEWMDAVYESVRRVWRH MGKQVRDWRKDVRSGERPKIRGYAKDVVGGNSIEQIEYLERQYKF LKSWSFFGKVSGQVIRAEKGSRFAITLREHIDHAKEDRLKKLADRIIM EALGYVYALDERGKGKWVAKYPPCQLILLEELSEYQFNNDRPPSEN NQLMQWSHRGVFQELINQAQVHDLLVGTMYAAFSSRFDARTGAP GIRCRRVPARCTQEHNPEPFPWWLNKFVVEHTLDACPLRADDLIPT GEGEIFVSPFSAEEGDFHQIHADLNAAQNLQQRLWSDFDISQIRLRC DWGEVDGELVLIPRLTGKRTADSYSNKVFYTNTGVTYYERERGKK RRKVFAQEKLSEEEAELLVEADEAREKSVVLMRDPSGIINRGNWTR QKEFWSMVNQRIEGYLVKQIRSRVPLQDSACENTGDI |
| AapCas12b | MAVKSMKVKLRLDNMPEIRAGLWKLHTEVNAGVRYYTEWLSLLRQ ENLYRRSPNGDGEQECYKTAEECKAELLERLRARQVENGHCGPA GSDDELLQLARQLYELLVPQAIGAKGDAQQIARKFLSPLADKDAVG GLGIAKAGNKPRWVRMREAGEPGWEEEKAKAEARKSTDRTADVL RALADFGLKPLMRVYTDSDMSSVQWKPLRKGQAVRTWDRDMFQQ AIERMMSWESWNQRVGEAYAKLVEQKSRFEQKNFVGQEHLVQLV NQLQQDMKEASHGLESKEQTAHYLTGRALRGSDKVFEKWEKLDP DAPFDLYDTEIKNVQRRNTRRFGSHDLFAKLAEPKYQALWREDASF LTRYAVYNSIVRKLNHAKMFATFTLPDATAHPIWTRFDKLGGNLHQY TFLFNEFGEGRHAIRFQKLLTVEDGVAKEVDDVTVPISMSAQLDDLL PRDPHELVALYFQDYGAEQHLAGEFGGAKIQYRRDQLNHLHARRG ARDVYLNLSVRVQSQSEARGERRPPYAAVFRLVGDNHRAFVHFDK LSDYLAEHPDDGKLGSEGLLSGLRVMSVDLGLRTSASISVFRVARK DELKPNSEGRVPFCFPIEGNENLVAVHERSQLLKLPGETESKDLRAI REERQRTLRQLRTQLAYLRLLVRCGSEDVGRRERSWAKLIEQPMD ANQMTPDWREAFEDELQKLKSLYGICGDREWTEAVYESVRRVWR HMGKQVRDWRKDVRSGERPKIRGYQKDVVGGNSIEQIEYLERQYK FLKSWSFFGKVSGQVIRAEKGSRFAITLREHIDHAKEDRLKKLADRII MEALGYVYALDDERGKGKWVAKYPPCQLILLEELSEYQFNNDRPP SENNQLMQWSHRGVFQELLNQAQVHDLLVGTMYAAFSSRFDART GAPGIRCRRVPARCAREQNPEPFPWWLNKFVAEHKLDGCPLRAD DLIPTGEGEFFVSPFSAEEGDFHQIHADLNAAQNLQRRLWSDFDIS QIRLRCDWGEVDGEPVLIPRTTGKRTADSYGNKVFYTKTGVTYYER ERGKKRRKVFAQEELSEEEAELLVEADEAREKSVVLMRDPSGIINR GDWTRQKEFWSMVNQRIEGYLVKQIRSRVRLQESACENTGDI |
| BrCas12b | MPVRSFKVKLVTRSGDAEHMLQLRRGLWKTHEIVNQGIAYYMNKL ALMRQEPYAGKSREVVRLELLHSLRAQQKRNNWTGDAGTDDEILN LSRRLYELLVPSAIGEKGDAQMLSRKFLSPLVDPNSEGGKGTAKSG RKPRWMKMREEGHPDWEAEREKDRAKKAADPTASILNDLEAFGLR PLFPLFTDEQKGIQWLPKQKRQFVRTFDRDMFQQALERMLSWES WNRRVAEEYQKLQAQRDELYAKYLADGGAWLEALQSFEKQREVE |

| | |
|---|---|
| | LAEESFAAKSEYLITRRQIRGWKQVYEKWSQLPEHAAQEQFWQVV<br>ADVQTSLPGAFGDPKVYQFLSQPEHHHIWRGYPNRLFHYSDYNGV<br>RKKLQRARHDATFTLPDPVEHPLWIRFDARGGNIHDYEISQNGKQY<br>QVTFSRLLWPENETWVERENVTVAIGASQQLKRQIRLDGYADKKQ<br>KVRYRDYSSGIELTGVLGGAKIQFDRRHLRKASNRLADGETGPVYL<br>NVVVDIEPFLAMRNGRLQTPIGQVLQVNTKDWPKVTGYKPAELISWI<br>QNSPLAVGTGVNTIEAGMRVMSVDLGQRSAAAVSIFEVMRQKPAE<br>QETKLFYPIAVTGLYAVHRRSLLLRLPGEKISDEIEQQRKIRAHARSL<br>VRYQIRLLADVLRLHTRGTAEQRRAKLDELLATLQTKQELDQKLWQ<br>TELEKLFDYIHEPAERWQQALVAAHRTLEPVIGQAVRHWRKSLRID<br>RKGLAGMSMWNIEELEETRKLLIAWSKHSRVPGEPNRLDKEETFAP<br>QQLQHIQNVKDDRLKQMANLLVMTALGYKYDEAEKQWKEAYPACQ<br>MILFEDLSRYRFALDRPRRENNRLMKWAHRSIPRLVYLQGELFGIQ<br>VGDVYSAYTSRFHAKTGAPGIRCHALKEEDLQPNSYVVKQLIKDGFI<br>REDQTGSLKPGQIVPWSGGELFVTLADRSGSRLAVIHADINAAQNL<br>QKRFWQQNTEIFRVPCKVTTSGLIPAYDKMKKLFGKGYFAKINQTD<br>TSEVYVWEHSAKMKGKTTPADPAEEGVFDESLTDEMEELEDSQEG<br>YKTLFRDPSGFFWSSDRWLPQKEFWFWVKRRIEKKLREQLQ |
